# Estimated Impact of Model-Guided Venous Thromboembolism Prophylaxis versus Physician Practice

**DOI:** 10.1101/2025.05.29.25328593

**Authors:** Benjamin G Mittman, Michael B Rothberg

**Affiliations:** Medical Scientist Training Program, School of Medicine, Case Western Reserve University, Cleveland, OH, USA; Center for Value-Based Care Research, Primary Care Institute, Cleveland Clinic, Cleveland, OH, USA; Department of Population and Quantitative Health Sciences, School of Medicine, Case Western Reserve University, Cleveland, OH, USA

**Keywords:** venous thromboembolism, hemorrhage, heparin, risk assessment, clinical utility, clinical decision support systems

## Abstract

**Background:** The American Society of Hematology (ASH) recommends assessing venous thromboembolism (VTE) and major bleeding risk to optimize pharmacological VTE prophylaxis for medical inpatients. However, the clinical utility of model-guided approaches remains unknown.

**Methods:** Our objective was to estimate differences in VTE and major bleeding event rates and efficiency with prophylaxis guided by risk models versus prophylaxis based on physician judgment. Patients were adults admitted to one of 10 Cleveland Clinic hospitals between December 2017 and January 2020. We compared physician practice with hypothetical prophylaxis recommended by model-based prophylaxis strategies, including ASH-recommended risk scores (Padua and IMPROVE) and locally derived Cleveland Clinic risk prediction models. For each strategy we quantified the prophylaxis rate, VTE and major bleeding rates, and the incremental number-needed-to-treat (NNT) to prevent one event (VTE or bleeding).

**Results:** Physicians prescribed prophylaxis to 62% of patients whereas model-based strategies recommended prophylaxis for 17-87%. Model-guided prophylaxis produced more VTEs and fewer major bleeds than physicians, but total events varied among strategies. Overall, per 1,000 patients, model-based strategies produced 14.0-16.1 events compared with 14.3 for physicians. The Padua/IMPROVE models recommended prophylaxis for the fewest patients but caused the most total events. The most efficient model-based strategy recommended prophylaxis to 28% of patients with an incremental NNT (relative to no prophylaxis) of 80. Compared to physicians, it reduced prophylaxis by 55% and total events by 0.14%.

**Conclusions:** Physicians often prescribed inappropriate prophylaxis, highlighting the need for decision support. A model-based strategy maximized efficiency, reducing both events and prophylaxis relative to physicians.

## Introduction

Venous thromboembolism (VTE) affects 300,000 to 600,000 people and causes up to 100,000 deaths each year in the United States.^1^ At least half of all cases are attributable to current or recent hospitalization.^2,3^ Multiple randomized controlled trials (RCTs) in medical inpatients have demonstrated that prophylactic anticoagulation reduces symptomatic VTE by 27% to 52% compared to placebo.^4–7^ However, anticoagulants carry risks of adverse effects; prophylactic doses increase major bleeding risk by an estimated 37% to 92%.^5–9^ Therefore, guidelines recommend assessing major bleeding risk and VTE risk concurrently to decide which patients should receive pharmacological prophylaxis.^6,8,10,11^ According to the guidelines, patients should receive prophylaxis if they are at high risk for VTE and acceptable risk for major bleeding.

Although some have argued in favor of simplified prescribing approaches that minimize the need for decision support,^12,13^ contemporary guidelines specifically endorse risk assessment for both VTE and bleeding. However, the real-world effects of model-based risk assessment are poorly understood because they have not been prospectively tested. Ideally, patients at low VTE risk would not receive prophylaxis, and those at high VTE risk would, if they were at low risk for major bleeding. These judgments are hard to make in clinical practice,^9,14^ but validated prediction models could help guide these decisions.^15–17^ The American Society of Hematology (ASH) guidelines recommend using the Padua^18^ and IMPROVE^19^ risk scores to assess VTE and major bleeding risk, respectively. Both models performed poorly in external validation studies^20,21^ and to date no one has assessed the clinical utility of concurrent usage of these models. In contrast, the Cleveland Clinic VTE Model^22^ (CCVM) and the Cleveland Clinic Bleeding Model^9^ (CCBM) have outperformed Padua and IMPROVE in the Cleveland Clinic population.

It remains unknown how either the ASH guideline-recommended models or the Cleveland Clinic models perform together, or whether either pair is better than physician judgment. To address this gap, we first characterized prophylaxis prescribing in a large cohort of medical inpatients. Next, we simulated prophylaxis according to guideline-recommended risk assessment models (Padua and IMPROVE) and the Cleveland Clinic models (CCVM and CCBM), as well as a strategy that simply avoided prophylaxis for patients at high risk of bleeding.^12^ We estimated hypothetical incidence rates of VTE and major bleeding and compared how efficiently different strategies minimized total events.

## Methods

### Setting and patients

Our cohort consisted of adults admitted to one of 10 Cleveland Clinic Health System (CCHS) hospitals between October 1, 2017 and January 31, 2020. Hospitals were located in Ohio and Florida and varied in size from a 126-bed community hospital to a 1,400-bed quaternary care academic medical center. We included patients aged 18 years or older who were admitted to a medical service, either directly or from the emergency room. For patients with multiple admissions during the study period, we randomly selected only one admission for inclusion. We excluded patients receiving therapeutic doses of anticoagulation because they would not be eligible for pharmacological VTE prophylaxis. All patient data were extracted from the Cleveland Clinic electronic health record (EHR) system and verified for accuracy and completeness. Statistical analyses were performed using R version 4.2.3.^23^ The study was approved by the Cleveland Clinic Institutional Review Board (IRB #14-240).

### Identification of venous thromboembolism

VTE was defined as upper and lower extremity deep vein thrombosis (DVT) or pulmonary embolism (PE) occurring within 14 days of hospitalization. VTE outcomes included events previously identified for the development and validation of the CCVM^22^ plus additional events identified for evaluation and updating of the current version of the model. Outcomes were identified using a combination of International Classification of Diseases (ICD) codes, diagnostic imaging tests, treatment records, and natural language processing, and verified by manual chart review.

### Identification of major bleeding

Major bleeding was assessed during hospitalization, using the definition from the International Society on Thrombosis and Hemostasis (ISTH)—clinically overt and either fatal or associated with one of the following: (a) fall in hemoglobin of 2 g/dL or more, (b) documented transfusion of at least 2 units of packed red blood cells, or (c) involvement of a critical anatomical site (e.g., intracranial, pericardial, intramuscular with compartment syndrome, retroperitoneal). Major bleeding outcomes for this cohort were previously identified in the study of the development and validation of the CCBM^9^ for major bleeding risk assessment. That study used a combination of ICD codes and laboratory values to identify major bleeds, all of which were verified by manual chart review.

### Identification of prophylaxis receipt

We identified receipt of pharmacological prophylaxis from the EHR’s medication administration record. Prophylaxis included unfractionated heparin (UFH; subcutaneous heparin 5,000 units 2–3 times daily), low molecular weight heparin (LMWH; enoxaparin 40mg daily or dalteparin 5,000 units daily), or a factor Xa inhibitor (fondaparinux 2.5mg daily) administered within 48 hours of admission.

### Characterizing physician practice

To understand how physicians’ prescribed prophylaxis related to patient risk, we stratified patients by VTE and major bleeding risk using the Cleveland Clinic (CC) models and calculated the percentage of patients that received prophylaxis within each risk quantile.

### Defining strategies for model-guided prophylaxis

We considered four possible prophylaxis strategies. Strategy 1 (Guidelines strategy) followed the American Society of Hematology (ASH) guidelines for pharmacological VTE prophylaxis in medical patients,^8^ which recommend prophylaxis for patients with a Padua score of ≥4 points,^18^ indicating high VTE risk, and an IMPROVE score <7 points,^19^ indicating low bleeding risk, and withholding prophylaxis from all other patients. Strategy 2 (CC Minimize-Bleeding strategy) used the Cleveland Clinic (CC) models to define high-risk, i.e., recommending prophylaxis for patients with a 14-day VTE risk ≥1%^24^ and an in-hospital bleeding risk <0.78%^9^. Strategy 3 (CC Minimize-Events strategy) also used the CC models but allowed prophylaxis for patients at high risk of both outcomes if their individual VTE risk outweighed their major bleeding risk. Strategy 4 (Near-Universal strategy) simplified prescribing by giving prophylaxis to all patients with low bleeding risk according to the CCBM, regardless of their VTE risk. We used the CCBM rather than the IMPROVE model because CCBM outperformed IMPROVE in this dataset.^9^

### Estimating clinical events

Our goal was to estimate how each strategy would affect rates of VTE and major bleeding compared with physicians. To do this, we first determined for each strategy which patients would receive prophylaxis. For patients indicated to receive prophylaxis, their probabilities of VTE and major bleeding risk as predicted by the CC models were adjusted by the relative risks of VTE efficacy and harm (e.g., in the base case, probability of VTE was reduced by 52% and probability of major bleeding was increased by 37%). If they did not receive prophylaxis, their predicted probabilities of VTE and major bleeding were the unadjusted CCVM and CCBM values. For each strategy, we summed all patients’ risks of events to get the total number of expected VTE and major bleeding events in the cohort. We divided each number of events by the size of the cohort and then multiplied it by 1,000 to get the rate per 1,000 individuals.

### Sensitivity analyses

There is moderate uncertainty regarding the efficacy and harm of pharmacological prophylaxis. Because these are key variables in our estimates, we evaluated the sensitivity of our primary outcomes to these variables, calculating the expected number of VTE and major bleeding events throughout the plausible ranges of efficacy (i.e., reduction in VTE) and harm (i.e., increase in major bleeding). Based on published studies, efficacy ranged from 27% to 52%^4–7^ reduction in VTE and harm ranged from 37% to 92% increase in major bleeding.^5–9^

### Comparing prophylaxis efficiency

There is a tension between the efficiency of prophylaxis and the goal of preventing events. More prophylaxis always reduces the incidence of VTE and increases the incidence of major bleeding, but prophylaxis is not equally efficient across risk groups. We compared how efficiently prophylaxis minimized total events with different prophylaxis strategies by calculating the additional number of patients that would need to receive prophylaxis to prevent one additional event, compared with the prior strategy. This is defined as the incremental number needed to treat (NNT), calculated according to the formula:

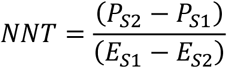

where *P* = the rate of prophylaxis per 100 eligible patients, *E* = the expected rate of total events per 100 individuals, [*S*_1_] denotes the baseline strategy, and [*S*_2_] denotes the comparator strategy. Thus, if the first strategy required prophylaxis for 20% of patients, and those patients had an average expected event rate of 1.6%, and the second strategy required prophylaxis for 50% of patients, and those patients had an average expected event rate of 1.3%, the NNT would be (50-20)/(1.6-1.3)=100.

## Results

### Patient characteristics

We identified 48,030 medical patients who were hospitalized during the study period. After excluding 2,005 (4.2%) patients who were missing one or more variables needed to determine VTE risk, major bleeding risk, or prophylaxis receipt, our final cohort contained 46,025 patients. Patients had a mean age of 61.4 years (SD = 19.0 years), 52.5% of patients were female, and 72.3% of patients were white. Patients’ 14-day VTE risk, computed by the CCVM, ranged from 0.31% - 45.3%, with a mean of 1.26% and a median of 0.74%. Probabilities of major bleeding, computed by the CCBM, ranged from 0.017% - 78.6%, with a mean of 0.52% and a median of 0.21%.

### Characterizing physician practice

A total of 28,396 (61.7%) patients received prophylaxis. Across VTE risk quantiles as computed by the CCVM, physician prophylaxis rates ranged from 50% to 87% (Figure 1). Physicians displayed modest sensitivity to VTE risk and almost no sensitivity to major bleeding risk. Overall, physicians treated >50% of low-risk patients and withheld prophylaxis from >20% of high-risk patients, according to the CCVM. Physicians prescribed prophylaxis to 66% of patients at highest risk for major bleeding (probability ≥10%) according to the CCBM.

**Figure 1.**
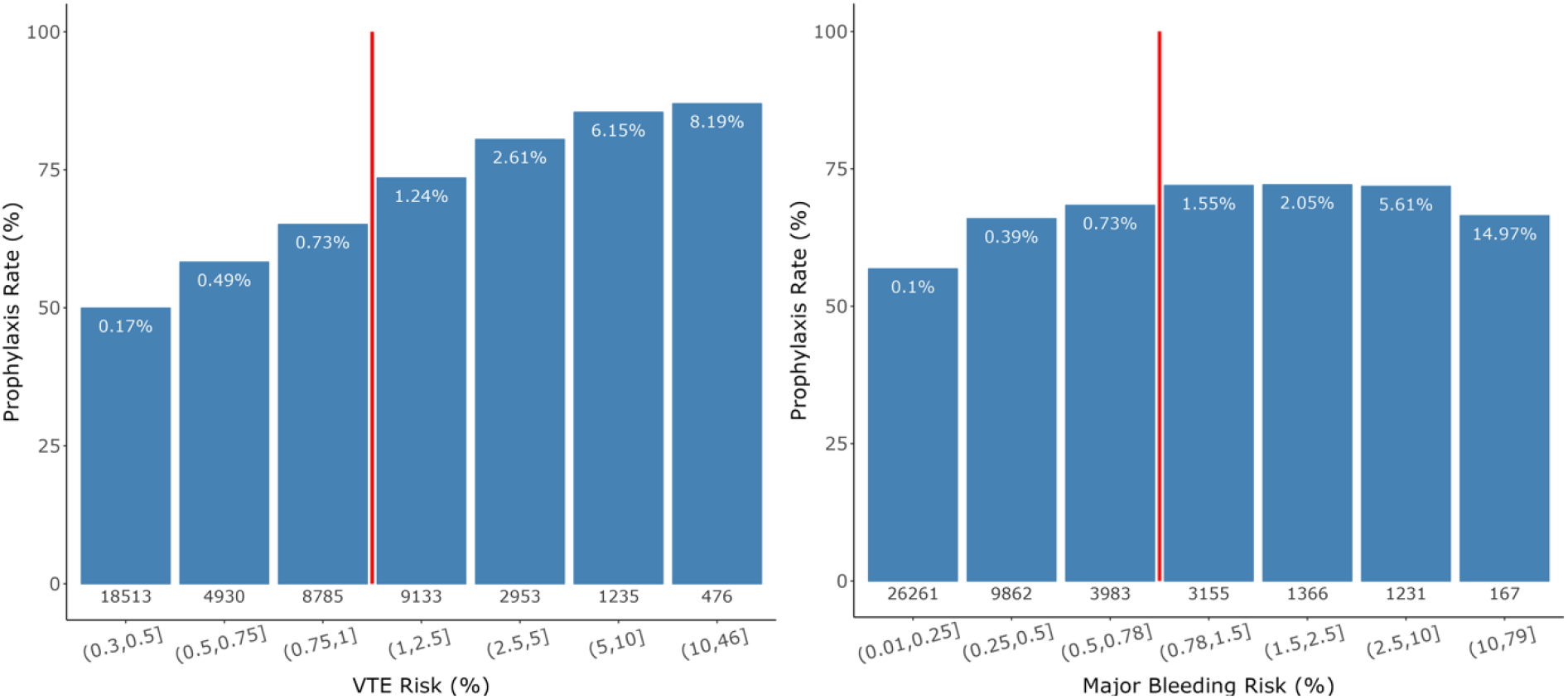
Observed prophylaxis rates stratified by VTE risk (left) and major bleeding risk (right). Risk probabilities were calculated using the Cleveland Clinic VTE model (CCVM) and Cleveland Clinic Bleeding model (CCBM). The value beneath each bar is the number of patients within that risk group. The percentage at the top of each bar is the observed rate of events within that risk group, demonstrating good concordance between actual and predicted risk for both models. Event rates are not adjusted for prophylaxis, which reduces risk of VTE and increases risk of major bleeding. As a result, predicted rates of VTE appear somewhat overestimated and risks of bleeding appear underestimated. The vertical red lines indicate the high-risk thresholds for the CCVM (1.0%) and CCBM (0.78%).

### Comparing physicians with model-guided prophylaxis

Figure 2 shows prophylaxis rates and clinical events for physicians and model-guided strategies. All the strategies had similar rates of events but drastically different rates of prophylaxis. The Near-Universal strategy had the highest prophylaxis rate and fewest events, while the guideline strategy had the lowest prophylaxis rate and the most events. The CC Minimize-Events strategy had approximately the same number of events as physicians, achieved with less than half the prophylaxis.

**Figure 2.**
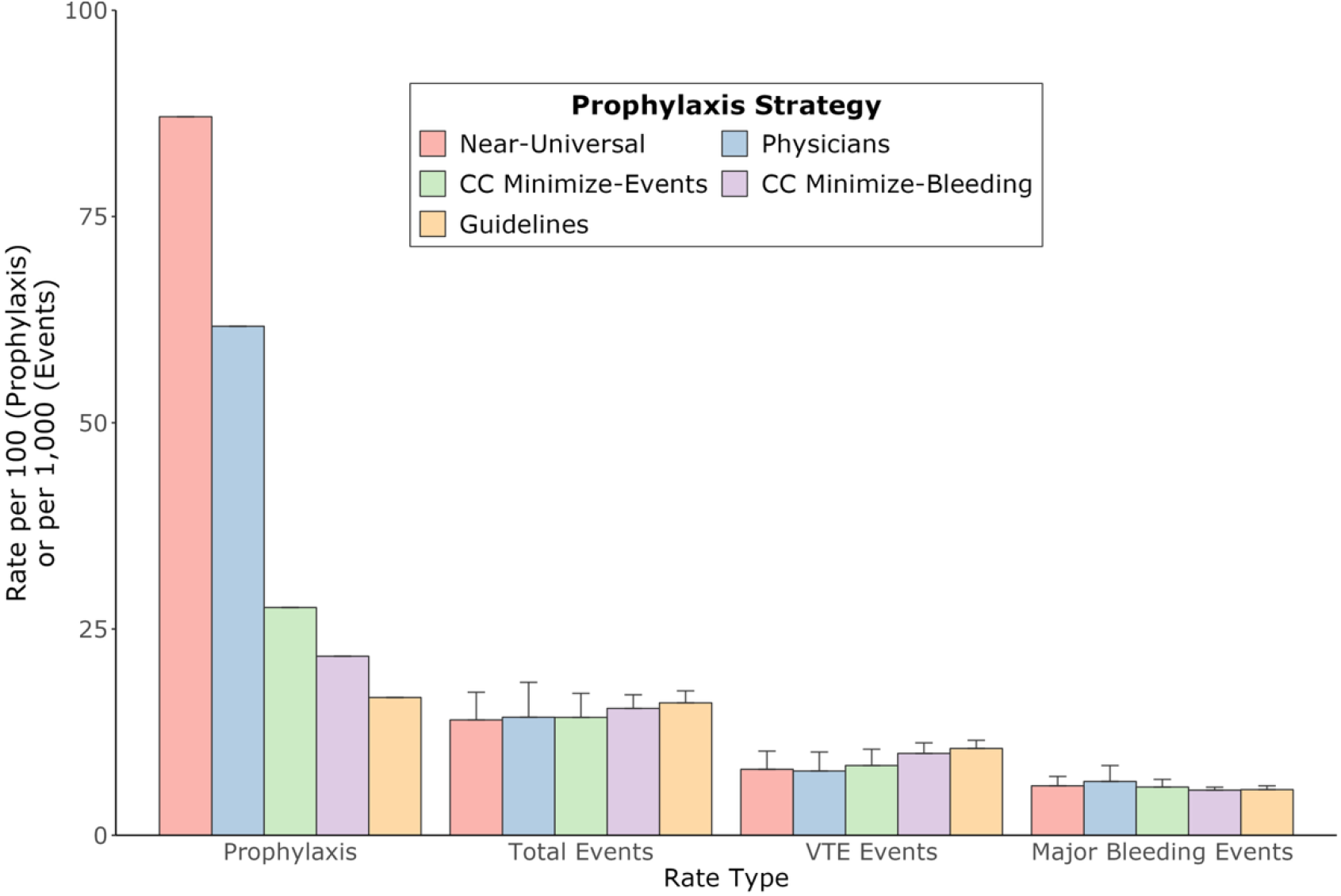
Prophylaxis and event rates for physicians (blue) versus four model-guided prophylaxis strategies: Cleveland Clinic (CC) models with prophylaxis for all patients regardless of VTE risk, except those with high major bleeding risk (pink); CC models with VTE and major bleeding equally balanced for high-risk patients (green); baseline CC models, with prophylaxis for high VTE risk and low major bleeding risk patients (purple); and guideline-recommended models, Padua and IMPROVE (orange). Bar heights are lower-bound event estimates based on high efficacy and high harm of prophylaxis; error bars are upper-bound estimates based on low efficacy and low harm.

### Comparing prophylaxis efficiency

Compared with no prophylaxis, the CC Minimize-Events strategy increased prophylaxis by 27.6% and reduced total events by 3.5 per 1,000, yielding an incremental NNT of 80 (Figure 3). The Near-Universal strategy required an additional 59.5% of patients to receive prophylaxis and further reduced total events by 0.3 per 1,000, for an incremental NNT of 1,920. All other strategies were considered “dominated;” that is, they used less prophylaxis but had more events per unit of prophylaxis than the CC Minimize-Events strategy. For example, compared to the CC Minimize-Events strategy, physicians prescribed 34.1% more prophylaxis and yet had slightly more total events, indicating net harm.

**Figure 3.**
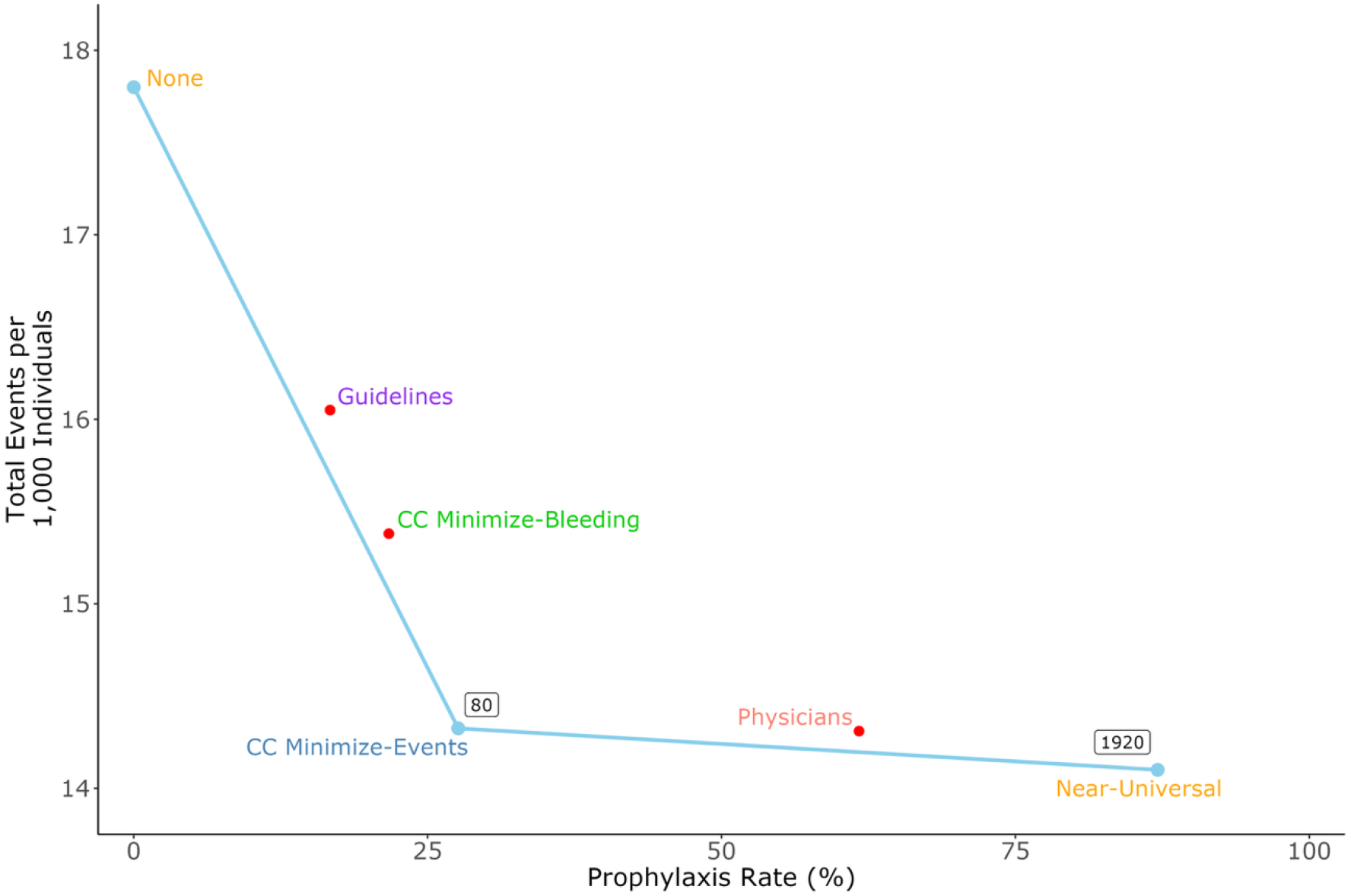
Efficiency curve showing prophylaxis rates versus total event rates for each prophylaxis strategy. Event rates shown are lower bound estimates from sensitivity analyses. Incremental NNTs (boxed values) are shown only for strategies along the efficiency frontier, i.e., the set of points with dominant efficiency.

### Sensitivity analyses

Each strategy produced a range of expected events and incremental NNTs due to uncertainty in the estimates of prophylaxis efficacy and harm (Table 1). For all combinations of prophylaxis efficacy and harm, the Near-Universal and CC Minimize-Events strategies always produced fewer events than physicians. The CC Minimize-Bleeding strategy (NNT = 105-280) dominated when prophylaxis harm was high, but the CC Minimize-Events strategy (NNT = 80-180) was similar to or more efficient than all other strategies in 3 of 4 analyses. Physicians and the Guidelines strategy did not exhibit dominant efficiency in any analysis. The Near-Universal strategy had a minimum NNT >1,000 and was never the most efficient strategy.

**Table 1.** Sensitivity of modeled event rates and numbers needed to treat for physician prophylaxis and model-guided strategies to varying estimates of prophylaxis harm and efficacy.

| Sensitivity Analysis Iteration | VTE Events per 1,000 | Major Bleeding Events per 1,000 | Total Events per 1,000 | Incremental NNT |
| --- | --- | --- | --- | --- |
| Low Efficacy (27%), Low Harm (37%) |  |  |  |  |
| Physicians | 10.09 | 6.52 | 16.61 | - |
| Guidelines | 11.51 | 5.53 | 17.04 | - |
| CC Minimize-Bleeding | 11.20 | 5.46 | 16.66 | - |
| CC Minimize-Events | 10.43 | 5.84 | 16.27 | 180 |
| Near-Universal | 10.20 | 5.99 | 16.19 | 7,440 |
| Low Efficacy (27%), High Harm (92%) |  |  |  |  |
| Physicians | 10.09 | 8.45 | 18.54 | - |
| Guidelines | 11.51 | 5.99 | 17.50 | - |
| CC Minimize-Bleeding | 11.20 | 5.83 | 17.03 | 280 |
| CC Minimize-Events | 10.43 | 6.77 | 17.20 | - |
| Near-Universal | 10.20 | 7.13 | 17.33 | - |
| High Efficacy (52%), Low Harm (37%) |  |  |  |  |
| Physicians | 7.79 | 6.52 | 14.31 | - |
| Guidelines | 10.52 | 5.53 | 16.05 | - |
| CC Minimize-Bleeding | 9.92 | 5.46 | 15.38 | - |
| CC Minimize-Events | 8.45 | 5.84 | 14.29 | 80 |
| Near-Universal | 7.99 | 5.99 | 13.98 | 1,920 |
| High Efficacy (52%), High Harm (92%) |  |  |  |  |
| Physicians | 7.79 | 8.45 | 16.24 | - |
| Guidelines | 10.52 | 5.99 | 16.51 | - |
| CC Minimize-Bleeding | 9.92 | 5.83 | 15.75 | 105 |
| CC Minimize-Events | 8.45 | 6.77 | 15.22 | 110 |
| Near-Universal | 7.99 | 7.13 | 15.12 | 5,950 |

Note: VTE, venous thromboembolism; NNT, number needed to treat; CC, Cleveland Clinic. Estimates of prophylaxis efficacy (decrease in VTE risk) and harm (increase in major bleeding risk) were identified from published literature. NNT values are only shown for strategies that exhibited dominant efficiency in the respective analysis. Overall, event rates and NNTs displayed modest sensitivity to varying estimates of prophylaxis efficacy and harm. The difference in total events between physicians and every other strategy overlapped with zero. However, the Guidelines strategy was most likely to increase total events compared with physicians, whereas the CC Minimize-Events strategy was most likely to reduce events. The Guidelines strategy and Physicians never exhibited dominant efficiency.

## Discussion

In this study, we identified four strategies for model-guided prophylaxis and quantified hypothetical rates of VTE plus major bleeding for each. We also characterized patterns of real-world, physician-prescribed prophylaxis in our cohort and compared it to model-guided prophylaxis. We found that when physicians prescribed according to their clinical judgment, inappropriate prophylaxis was common, including overuse in >50% of low-risk patients and underuse in >20% of high-risk patients, consistent with other studies.^25,26^ Compared to physicians, the model-based strategies would reduce prophylaxis by at least half and up to nearly three-quarters without substantially increasing total expected events. These findings emphasize the need for clinical decision support using validated risk assessment models.

Although the models all outperformed the physicians, they were not all equally good. Compared to the physicians, the ASH guidelines, which used the Padua and IMPROVE risk scores, reduced prophylaxis the most, but at the cost of additional VTEs. The Cleveland Clinic models were more efficient, reducing prophylaxis in a more targeted way, so that VTEs were not increased relative to physicians. The most efficient approach used the Cleveland Clinic models to weigh the risks of VTE and major bleeding and prescribe prophylaxis whenever the former exceeded the latter in patients with high VTE risk. Guiding VTE prophylaxis in this way, using the most accurate models and optimal probability thresholds,^27^ could standardize practice across physicians and hospitals and improve risk communication between physicians and patients.^28,29^ This is an important distinction, because current guidelines emphasize not prescribing to patients at high risk of bleeding, even though many of them are at even higher risk of VTE.

In recommending prophylaxis, guidelines have not explicitly considered efficiency, and some authors have suggested a simplified approach of universal prophylaxis so long as patients are not at high risk of bleeding.^12^ The Near-Universal strategy, which does just that, did indeed minimize total events. However, compared to the CC Minimize-Events strategy, it required prophylaxing almost 2000 patients to prevent one additional event, a number not likely acceptable to physicians. In fact, in a survey of more than 200 hospitalists, when physicians were asked “What is the largest number of patients that you would be willing to give prophylaxis to in order to prevent one VTE?”, ^27^ the mean response was 87, and only one physician said more than 1000. This suggests that the CC Minimize-Events strategy would be acceptable to most physicians, whereas the Near-Universal strategy would not.

This study is to our knowledge the first to directly compare the clinical utility of model-guided prophylaxis with physician practice using real-world data. Many studies have described patterns of physician practice and highlighted the need for decision support. However, most available risk scores have shown poor performance in external validation studies,^30,31^ in some cases offering no discernible utility.^20^ In an attempt to determine the optimal prophylaxis approach, one study converted the IMPROVE risk scores for VTE and bleeding into a fast-and-frugal decision tree (FFT), showing that it could reduce unnecessary prophylaxis and lower costs.^32^ However, that study considered only the IMPROVE risk scores, whereas the Padua risk score is widely used for VTE risk assessment and recommended by the ASH guidelines.^8^ More recently, the Cleveland Clinic VTE^22^ and bleeding^9^ prediction models have outperformed both Padua and IMPROVE and offer the distinct advantage of predicting patient risk in terms of probability rather than a semi-quantitative point-based system, thereby allowing a direct comparison of the two risks. Our work expands on the FFT study by considering several model-guided prophylaxis strategies with different decision support tools and comparing both clinical outcomes and efficiency of treatment. Our study provides the strongest evidence yet that model-guided prophylaxis could safely reduce unnecessary prophylaxis.

Our findings should be considered in the context of this study’s limitations. These include our use of patient data from a single health system, which limits generalizability. We did not consider heparin-induced thrombocytopenia (HIT), although it is very rare with a prevalence <0.1%^33^ and therefore less important than major bleeding. Our event modeling made important assumptions that should be revisited in future work. We applied the same estimates of prophylaxis efficacy and harm to all patients, but heparin may exhibit different properties in different patients.^34^ Follow-up studies should investigate whether prophylaxis affects VTE and major bleeding risk differently for different subpopulations or risk groups. We also assumed that a VTE or a major bleed is equally harmful regardless of the patient. The dynamics of recurrence, long-term sequelae, and differences between symptomatic and asymptomatic VTE could influence how physicians decide to prioritize competing VTE and major bleeding risks in different patients.^35,36^ We adopted a global approach that assumes these differences are balanced at the population level, but future work should compare our approach among different subgroups of medical patients. Lastly, we calculated each patient’s risk based on the Cleveland Clinic models. To the extent that they are miscalibrated, our findings would be different. However, calibration in the Cleveland Clinic data is excellent.^9,22^

## Conclusion

We compared the clinical implications of physician practice for pharmacological VTE prophylaxis with multiple model-guided strategies. Each strategy increased VTEs but reduced major bleeds but there was a wide range of prophylaxis rates. Using the Padua and IMPROVE risk scores, as recommended by the ASH guidelines, would minimize prophylaxis but increase total events. On the other hand, using the Cleveland Clinic models would most efficiently minimize total events and reduce prophylaxis.

## Data Availability

De-identified data will be shared by request on a case-by-case basis, consistent with Cleveland Clinic data sharing policies.

## Author Contributions

BGM and MBR contributed to study conception, design, data analysis, and interpretation. BGM drafted the manuscript under the supervision of MBR.

